# Extending the range of symptoms in a Bayesian Network for the Predictive Diagnosis of COVID-19

**DOI:** 10.1101/2020.10.22.20217554

**Authors:** Rachel Butcher, Norman Fenton

**Affiliations:** Queen Mary University of London

**Keywords:** SARS-CoV-2, COVID-19, Bayesian network, predictive model

## Abstract

Emerging digital technologies have taken an unprecedented position at the forefront of COVID-19 management. This paper extends a previous Bayesian network designed to predict the probability of COVID-19 infection, based on a patient’s profile. The structure and prior probabilities have been amalgamated from the knowledge of peer-reviewed articles. The network accounts for demographics, behaviours and symptoms, and can mathematically identify multivariate combinations with the highest risk. Potential applications include patient triage in healthcare systems or embedded software for contact-tracing apps. Specifically, this paper extends the set of symptoms that are a marker for COVID-19 infection and the differential diagnosis of other conditions with similar presentations.

## I. Introduction

In January 2020, the UK welcomed a new decade, unknowingly about to face its second pandemic of the 21^st^ century. By late 2019, a novel strain of coronavirus had made the zoonotic leap from an unknown animal reservoir to humans, in connection with a seafood market in Wuhan, China (WHO, 2020b). The virus (hereinafter ‘SARS-CoV-2’) is homologous to that of the 2002-2003 SARS epidemic, causing asymptomatic to severe respiratory disease (hereinafter ‘COVID-19’) (Yuki *et al*., 2020). The virulent infection since reached 213 countries and territories globally. To date, confirmed cases exceed 19 million with over 700,000 reported deaths (worldometers, 2020). On 11^th^ March 2020, the WHO promoted COVID-19 to pandemic status.

The pandemic has been a huge burden on the global economy. In the absence of a vaccine, public health response has been targeted towards disease containment (Wynants *et al*., 2020). On 23^rd^ March 2020, the UK was placed under strict lockdown in attempts “flatten the curve”. Such measures aimed to maintain the number of cases below healthcare system capacity (Matrajt and Leung, 2020). Widespread lockdown forced closure of much of the retail, hospitality, and entertainment sectors. From May to June, the UK’s gross domestic product (GDP) dropped by 20.4%, marking the entrance into technical recession (Office for National Statistics, 2020a). Individuals in such sectors have been disproportionally affected, with unemployment reaching 3.6% and expected to rise (Office for National Statistics, 2020b). Further detrimental impacts have been felt across education, travel (Nicola *et al*., 2020) and mental health (Liang *et al*., 2020).

Despite a desperate need to bolster the economy, easing of restrictions carry the risk of disease resurgence and unsustainable strain on the NHS (Anderson, 2020). Whilst transmission surveillance remains an integral part of government response, community diagnosis will continue to be a limiting factor. Clinical tests, such as reverse transcription polymerase chain reaction (RT-PCR), are hindered by significant false-negative rates (West *et al*., 2020), expensive equipment and the need for specialised professionals (Russo *et al*., 2020). The management of COVID-19 has seen an unprecedented dependence on emerging digital technologies. AI-driven predictive models have the potential to provide an immediate diagnosis based on a patient’s profile, without the need for testing (Wynants *et al*., 2020). Technologies can be used to reduced triage time in hospitals (Soltan *et al*., 2020) or improve community outreach where testing is inaccessible.

This study builds on the Bayesian network solution by Fenton *et al*., (2020) and concurrent work by Prodhan (2020) for prediction of current COVID-19 status coupled with eventual prognoses. The network takes an input of observable symptoms and risk factors to produce a personalised probability score for disease status.

## II. Related Work

### A. Bayesian Networks

Bayesian networks (BNs) exploit Bayes probabilistic reasoning to provide insights in the causal relationships between the contributors and outcomes of an event (He, 2014). The BNs ability to account for uncertainties makes it a useful application to support clinical decision-making, where the event is the probability of disease in a patient (Wang *et al*., 2014). The network graphically represents the directed conditional dependencies (arcs) between stochastic variables (nodes) of the event. Given node A represents infection with a disease, e.g. flu, and node B represents some symptom, e.g. cough, the directed arc would point from A to B, and be interpreted as A causes or influences B. A may also be expressed as the parent of B (Fenton and Neil, 2018). Associated with each variable is a predefined set of mutually exclusive states and a node probability table (NPT) (Jensen, 1996). The NPT defines the marginal probability distribution across the states. When an observation is set for a known variable, the effect propagates both forwards and backwards to update the posterior probability of any dependencies. Important constraints of a BN is it must be acyclic, as the algorithm does not handle feedback loops, and it should not be a complete graph, meaning variables independent of each other should not share an arc (Fenton and Neil, 2018).

BNs are credible in decision support as they can provide evidence of the reasoning behind their conclusions. Physicians can simply trace the posterior probabilities through the network interface or use secondary tools such as BANTER (Kahn *et al*., 1997). BANTER identifies the most influential nodes using sensitivity analysis and the strongest path of influence to the hypothesis. A textual explanation is then generated for the physician to review (Haddawy *et al*., 1994). A study by Wang *et al*., (2014) proposed a BN model for comparison against traditional machine learning methods, namely logistic regression, naive Bayes and support vector machine, for the prediction of lung cancer-induced brain metastasis. Whilst sensitivity was only marginally improved by the BN, the method’s major advantages were realised as: easy comprehension, efficient modelling of both linear and non-linear events, the ability to reason from consequence to cause and the ability to handle missing data. Bayes theorem provides a means to standardise the interactions between dependent and independent probabilities, allowing for reasoning under uncertainty and incorporation of domain knowledge (Kahn *et al*., 1997). Uncertainties are missing observations, such as unavailable information on patient family history or diagnostic tests that have not yet been performed. BNs are advantageous as they can maintain accurate predictions even with little observational input (Zheng *et al*., 2008). Statistical paradoxes are prevalent in literature and can drive false claims in the media or, more critically, incorrect conclusions in medical studies. Simpson’s paradox occurs when conclusions derived from aggregated data reverse when the same data are stratified The common clinical trial example cited is; when two treatments A and B are administered to a population of patients, treatment A has greater efficiency. However, when the same population is stratified, for example by sex, treatment B paradoxically performs better in each sub-category. Given an assumption about causality, BNs can resolve such paradoxes by computing the necessary, but often counterintuitive, inverse probabilities and simulate the effect of an intervention, or ‘treatment’ in the given scenario (Fenton and Neill, 2019).

Numerous Bayesian models have been published since their popularisation by Judea Pearl in the late eighties (Pearl, 1988). Seixas *et al*., (2014) developed a network to support dementia and Alzheimer’s diagnosis using predisposal factors, neurophysiological test results, demographics and symptoms, populated by both expert knowledge and supervised learning. The BN was found to outperform other classifiers, such as decision tables, in diagnostic accuracy. Luciani *et al*., (2003) explored BNs diagnostic ability for pulmonary embolism, with nodes representing patient risk factors and pathophysiology, and NPTs populated by a systematic review of literature. A study by Kahn *et al*., (1997) constructed a BN to aid in the interpretation of mammographs for diagnosis of breast cancer. The model incorporates patient histories, physical symptoms, and mammographic indicators. Probabilities were mined from medical literature and expert mammographers. The network was successful in the early detection of breast cancer, allowing for pre-emptive medical intervention and ultimately improving patient prognosis. In 2001, Kahn *et al*., tailored another BN towards diagnosis of primary bone tumours, with nodes representing patient demographics, physical findings and lesion properties. The NPT values were elicited from peer-reviewed literature and the model returned a 68% diagnostic accuracy. BNs have demonstrated their value not only in medicine, but also agriculture. Bi and Chen (2011) developed a model for producing BNs for the diagnosis of crop diseases, such as corn borer which can reduce corn harvest by up to 15% per year and have significant economic damage.

Whilst BNs for diagnostic decision support is not a novel application, there has been poor uptake in clinical practice thought to be partially due to concerns about accuracy and a lack of evidence supporting clinical credibility (Yet *et al*., 2017).

### B. Predictive Models for COVID-19 Diagnosis

Several predictive models have recently emerged to help alleviate the burden of COVID-19 on healthcare systems. Such models may diagnose current infection, predict risk of infection or prognose disease progression to inform medical decision-making (Wynants *et al*., 2020). A systematic review of COVID-19 prediction models by Wynants *et al*., (2020) found current models to claim moderate to excellent predictive performance yet are severely limited by overfitting and bias. As adequate data may not always be available, it is suggested that future models comprehensively describe the demographics of the population on which the model is developed. Performance of the model can then be evaluated in relation to its applicability to the future user. Wynants *et al*., (2020) also propose basing the model on global rather than local patient data, to allow for greater application and generalisability. Both recommendations will be considered in the development of our Bayesian network.

Soltan *et al*., (2020) of Oxford University have developed two AI-driven models for triage of hospitalized, potential COVID-19 patients awaiting RT-PCR screening. Machine learning methods (namely logistic regression, random forest and extreme gradient boosted trees) were applied to electronic health record data from Oxford University Hospitals. Prediction was based on clinical data typically available for patients presenting to hospital, such as leukocyte counts and respiratory rate. Both models, all presenting patients and only admitted patients, achieved high sensitivity and specificity, and could reduce triage time from up to ∼48 hours (for RT-PCR) to ∼1 hour (for blood and vitals collection). However, the method is limited to individuals with symptoms severe enough to present to hospital and the input data requires specialist equipment and trained professionals. The models are currently in clinical trials under the name CURIAL AI, ultimately for use by the NHS.

Menni *et al*., (2020) of King’s College London studied potential symptoms predictive of COVID-19 infection. Using self-reported symptoms of the ZOE Global smartphone app, logistic regression determined anosmia, fever, persistent cough, fatigue, shortness of breath, diarrhea, delirium, skipped meals, abdominal pain, chest pain and hoarse voice were associated with a positive SARS-CoV-2 test in UK participants. Whilst the study incorporates a significantly large population and adjusts for age, sex and BMI; Menni *et al*., (2020) admit that the self-reported nature of the data is a caveat to the study being representative of the whole population. Symptoms have not been clinically verified, leaving opportunity for human error, and the sample population is reflective only of self-selected participants using the app and laboratory tested positive for SARS-CoV-2. Similarly, the study only queries known symptoms of COVID-19 and does not contribute to discovery of symptoms based on empirical evidence.

## III. Materials and Methods

### A. Requirements

The aim was to develop a probabilistic causal model for the prediction of COVID-19 infection and progression in an individual. To provide an accurate diagnosis, the model must account for all variables that may contribute to probability of infection (e.g. occupation, ethnicity, health risks or other demographics). As SARS-CoV-2 is thought to be transmitted by droplet infection, behaviours that increase human proximity and contact are also deemed high risk (ECDC, 2020a). The model should mine expert domain knowledge from peer-reviewed research to develop informed prior probabilities. The knowledge-based structure should also adhere to Bayesian network constraints. Rationale for the model, beyond what is discussed in this paper, is presented by Fenton et al., (2020). See Fig. 2. for complete model structure.

**Fig. 1.**
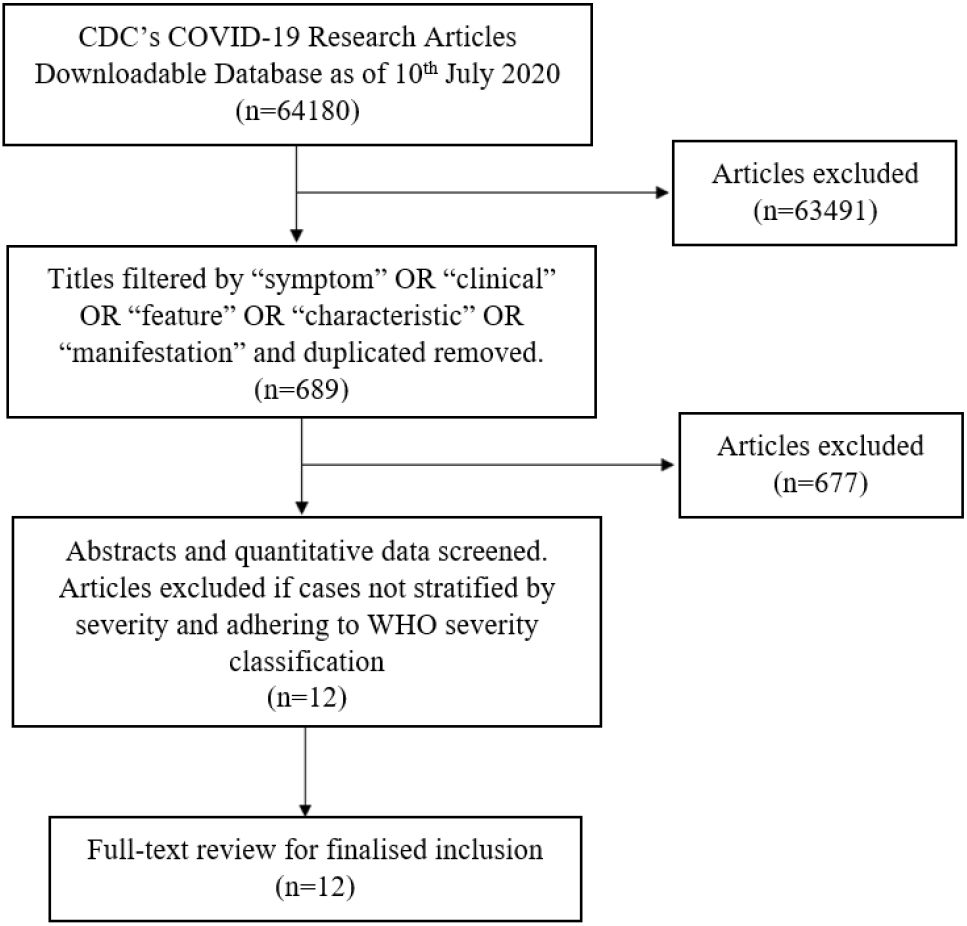
Flowchart describing the inclusion criteria for the systematic review.

**Fig. 2.**
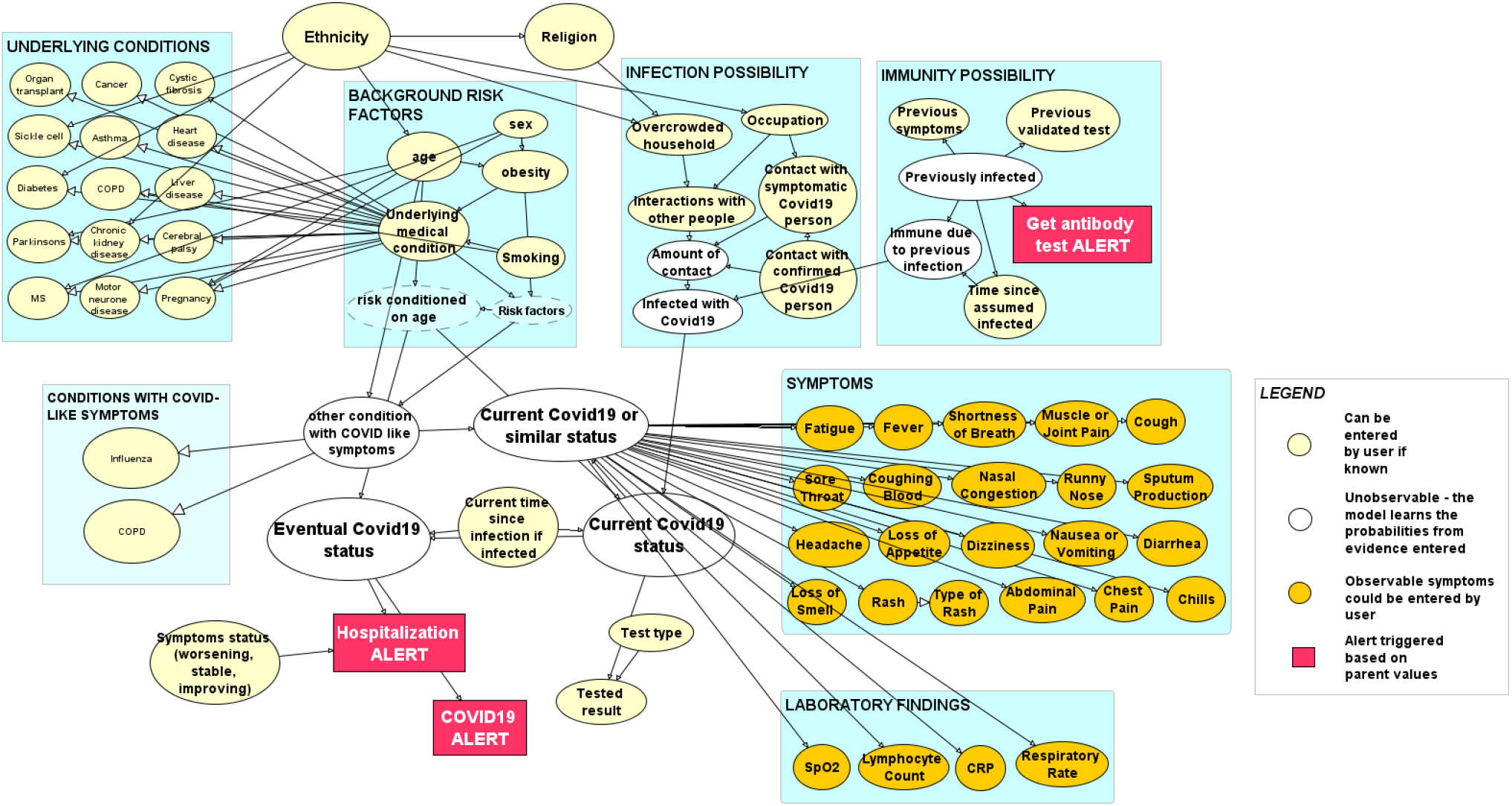
Complete structure of proposed Bayesian network..

### B. Design

As the model was a collaborative effort, this paper focuses primarily on symptom discovery and the differential diagnosis of conditions with similar clinical manifestations. Symptoms were mined on an empirical basis. Inclusion was not limited to the three main symptoms cited by the NHS: high temperature, continuous cough or a loss of smell or taste (NHS, 2020). A systematic review mitigates the constraints of a targeted search, revealing several additional symptoms that may improve the model’s sensitivity. The model can also afford to be dynamic, updating as new information comes to light.

Evaluating multiple presentations of COVID-19 was deemed an important step in model development. The novelty of COVID-19 means symptoms are not yet fully defined. Those that are commonly expressed, such as fever and coughing, are extremely non-specific and could point to a number of different diagnoses. To establish how COVID-19 presented in individuals, a systematic review was conducted using the CDC’s COVID-19 Research Articles Downloadable Database (Centers for Disease Control and Prevention, 2020). The database is updated daily with all publicly available COVID-19 research, including preprints, amalgamated from the following publishers: Medline, PubMed Central, Embase, CAB Abstracts, Global Health, PsycInfo, Cochrane Library, Scopus, Academic Search Complete, Africa Wide Information, CINAHL, ProQuest Central, SciFinder, the Virtual Health Library, LitCovid, WHO COVID-19 website, CDC COVID-19 website, Eurosurveillance, China CDC Weekly, Homeland Security Digital Library, ClinicalTrials.gov, bioRxiv, medRxiv, chemRxiv, and SSRN (Centers for Disease Control and Prevention, 2020).

The database was captured on the 10^th^ July 2020 and initially consisted of 64180 unique studies. Results were primarily screened for titles containing one, or a combination, of the terms: “symptom” OR “clinical” OR “feature” OR “characteristic” OR “manifestation”. On removal of duplicates, 689 unique titles remained. Each remaining study was screened by abstract and quality of quantitative data available. The inclusion criteria were met if: (a) quantitative symptom data was stratified by disease severity, (b) severe COVID-19 infection was defined by the WHO guidelines (WHO, 2020a), (c) cases were confirmed by a positive RT-PCR test. It is recognised that using RT-PCR alone is a caveat due to high false-negative reports, but it allowed for a standardized method of comparing symptoms. 12 papers satisfied the criteria and were used in development (Fig. 1.). It should also be noted that all studies were in English and preprint papers were not excluded from eligibility.

The research detailed 8095 cases, reported between 11th December 2019 to 20^th^ April 2020. Locations include Henan, Zhejiang and Wuhan in China, California, Spain, Tokyo, and Daegu in Korea; with a total of 4062 male patients and 4033 female patients (Table. 1). Frequency analysis was used to determine the most reported symptoms. Typically, if a symptom was found in two or more studies it was eligible for inclusion. The frequency target was low (2+) to mitigate potential effects of researcher bias or expectation, or effects of increased media attention on symptom reporting (Menni *et al*., 2020).

**TABLE I.**
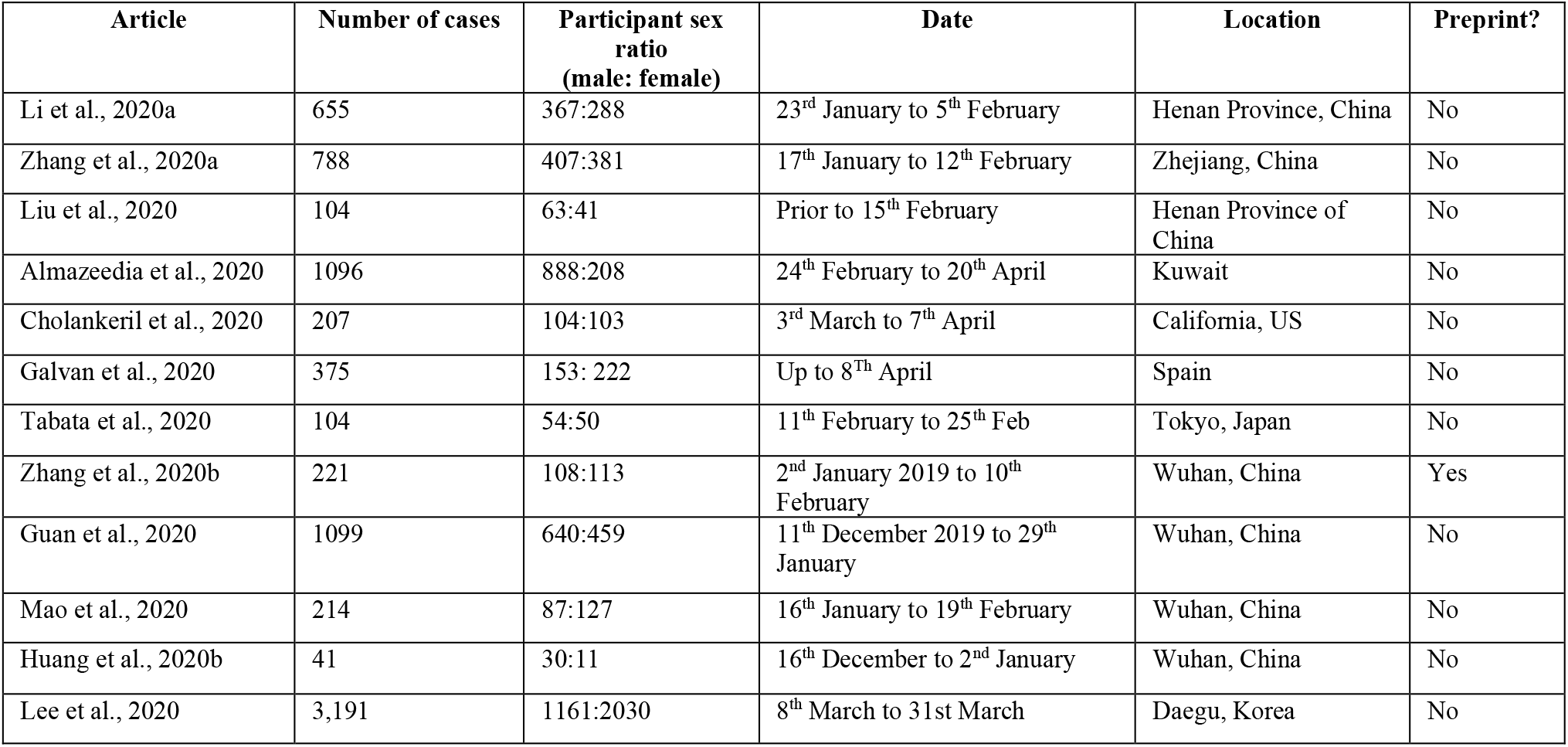
Table displaying features of the final articles selected by systematic review.

Each symptom identified by frequency analysis was cross-referenced with its likelihood ratio (LR) from Wagner *et al*., (2020a) curation of clinical notes (Table. 2). The likelihood ratio was calculated as follows (1)

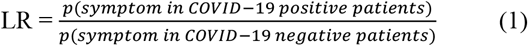

**TABLE II.** Table displaying the likelihood ratios of symptoms in indicating covid-19 infection (wagner et al., 2020a).

| Symptom | Likelihood Ratio |
| --- | --- |
| Anosmia or Ageusia | 27.08 |
| Fever/Chills | 2.57 |
| Cough | 2.24 |
| Rhinitis | 2.19 |
| Respiratory Difficulty | 2.18 |
| Myalgia/Arthralgia | 2.01 |
| Congestion | 1.73 |
| Headache | 1.71 |
| Hemoptysis | 1.48 |
| Pharyngitis | 1.42 |
| Change in appetite | 1.35 |
| Diarrhea | 1.35 |
| Fatigue | 0.89 |
| Chest pain/pressure | 0.78 |
| Gastrointestinal | 0.59 |
| Wheezing | 0.42 |
| Dermatitis | 0.33 |

Cells shaded grey indicate symptoms with a likelihood ratio greater than 1.

A likelihood score >1 is generally accepted to indicate that the symptom is a predictor for disease (McGee, 2002). Most discovered symptoms adhered to a result >1, with the exception of fatigue, chest pain, dermatitis and generalised gastrointestinal symptoms. However, these symptoms appeared frequently in literature and therefore deemed too indictive to omit. The strongest score was from anosmia (27.08), which is reflected in several other studies (Menni *et al*., 2020; Williams *et al*., 2020a).

The model has been designed to capture laboratory findings that explicitly define severe COVID-19 under WHO guidelines. An observation of respiratory rate ≥30 breaths per minute or oxygen saturation of <90% should calculate an increased prior probability of severe disease compared to mild (WHO, 2020a). Multiple studies also suggest elevated serum C-reactive protein (CRP) and lymphocytopenia provide important markers for disease severity (Ali, 2020; Wang, 2020; Chen *et al*., 2020a; Wagner *et al*., 2020b and Huang and Pranata, 2020a). Nodes reflecting this have been added as appropriate.

The differential diagnosis of COVID-19 is important in clinical decision-making. Clinical presentations of COVID-19 are often non-specific and can lead to misdiagnosis. The model considers the prevalence of other diseases with similar symptoms in the UK, as conditioned on both age and level of risk. Such conditions are often respiratory diseases; commonly cited misdiagnoses being influenza, chronic obstructive pulmonary disease (COPD) and meningitis. If it is observed prior to COVID-19 diagnosis that the subject suffers from any of these, then the model can account for such scenario and predict the probability of co-infection.

### C. Implementation

Specialised Bayesian network software AGENARISK has been used to host the model. AGENARISK automatically produces the format for NPTs and handles Bayes calculations when run.

Quantitative data from the systematic review was used to complete the NPTs. As children nodes of ‘Current COVID-19 or Similar’, each symptom required prior probabilities for: mild disease, severe disease, asymptomatic disease, non-COVID-19 similar disease and no disease. Where possible, data from multiple studies of the systematic review were amalgamated to produce robust prior probabilities. Data was only pooled across studies if the parameters were standardized. For example, fever was ubiquitous in all studies, but figures were only selected if it had been defined as >37.3°C. See Table. 3 for full pooled analysis by symptom. Whilst it is expected that asymptomatic disease would produce no symptoms, an estimated probability of 0.01 was included under the assumption that individuals categorised as such may be subclinical (Hu *et al*., 2020). Data for conditions with similar symptoms were gathered from Chen *et al*., (2020b), Menni *et al*., (2020) and Zayet *et al*., (2020), as part of a targeted literature search. SARS-CoV-2 negative RT-PCR results in patients presenting to hospital with symptoms were deemed an accurate prediction for the presence of a different disease. Such studies also looked at differences in symptom prevalence in COVID-19 vs non-COVID-19 pneumonia, and influenza. The probabilities of symptoms in subjects with no disease were taken from the REACT study by Riley *et al*., (2020). It has been assumed that symptoms reported by participants testing negative for COVID-19 were circumstantial since the study was community outreach rather than in a clinical setting. Type of Rash data was taken from studies by Zhao *et al*., (2020) and Matar *et al*., (2020), whilst laboratory findings were mined from Zhang *et al*., (2020b); Tabata *et al*., (2020); Guan *et al*., (2020); de Jager *et al*., (2010); Wilkerson *et al*., (2020) and J. Xie *et al*., (2020).

**TABLE III.** Table displaying the reference papers for each symptom with the combined number of observations over the combined number of participants

| Symptom | References | Pooled data: mild | Pooled data: severe |
| --- | --- | --- | --- |
| Fever | Li et al., 2020a<br>Zhang et al., 2020a<br>Zhang et al., 2020b<br>Guan et al., 2020<br>Huang et al., 2020b | 1997/2413 | 367/445 |
| Cough | Li et al., 2020a<br>Zhang et al., 2020a<br>Liu et al., 2020 | 1923/3819 | 462/677 |
|  | Almazeedia et al., 2020<br>Cholankeril et al., 2020<br>Tabata et al., 2020<br>Zhang et al., 2020b<br>Guan et al., 2020<br>Mao et al., 2020<br>Huang et al., 2020b |  |  |
| Fatigue | Li et al., 2020a<br>Zhang et al., 2020a<br>Liu et al., 2020<br>Almazeedia et al., 2020<br>Tabata et al., 2020<br>Zhang et al., 2020b<br>Guan et al., 2020<br>Huang et al., 2020b | 822/3584 | 272/491 |
| Sputum Production | Li et al., 2020a<br>Zhang et al., 2020a<br>Liu et al., 2020<br>Almazeedia et al., 2020<br>Guan et al., 2020<br>Huang et al., 2020b | 727/3373 | 145/408 |
| Chest Pain | Li et al., 2020a<br>Zhang et al., 2020a | 78/657 | 27/102 |
| Headache | Li et al., 2020a<br>Zhang et al., 2020a<br>Liu et al., 2020<br>Almazeedia et al., 2020<br>Tabata et al., 2020<br>Zhang et al., 2020b<br>Guan et al., 2020<br>Mao et al., 2020<br>Huang et al., 2020b | 372/3666 | 71/617 |
| Muscle or Joint Pain | Li et al., 2020a<br>Zhang et al., 2020a<br>Liu et al., 2020<br>Almazeedia et al., 2020<br>Cholankeril et al., 2020<br>Guan et al., 2020 | 552/4420 | 142/628 |
| Sore Throat | Li et al., 2020a<br>Zhang et al., 2020a<br>Liu et al., 2020<br>Almazeedia et al., 2020<br>Cholankeril et al., 2020<br>Tabata et al., 2020<br>Zhang et al., 2020b<br>Guan et al., 2020<br>Mao et al., 2020 | 503/3791 | 92/664 |
| Runny Nose | Li et al., 2020a<br>Liu et al., 2020<br>Tabata et al., 2020 | 52/609 | 14/130 |
| Shortness of Breath | Li et al., 2020a<br>Liu et al., 2020<br>Almazeedia et al., 2020<br>Cholankeril et al., 2020<br>Tabata et al., 2020<br>Zhang et al., 2020b<br>Guan et al., 2020<br>Huang et al., 2020b | 279/3020 | 199/473 |
| Diarrhea | Li et al., 2020a<br>Zhang et al., 2020a<br>Liu et al., 2020<br>Almazeedia et al., 2020<br>Cholankeril et al., 2020<br>Galvan et al., 2020<br>Tabata et al., 2020<br>Zhang et al., 2020b<br>Guan et al., 2020<br>Mao et al., 2020<br>Huang et al., 2020b<br>Lee et al., 2020 | 126/3107 | 70/599 |
| Nasal Congestion | Li et al., 2020a<br>Liu et al., 2020<br>Almazeedia et al., 2020<br>Guan et al., 2020 | 107/2637 | 10/317 |
| Nausea or Vomiting | Li et al., 2020a<br>Almazeedia et al., 2020<br>Cholankeril et al., 2020 | 110/3293 | 26/419 |
|  | Guan et al., 2020 |  |  |
| Coughing Blood | Zhang et al., 2020a<br>Almazeedia et al., 2020<br>Guan et al., 2020<br>Huang et al., 2020b | 16/2716 | 12/306 |
| Dizziness | Liu et al., 2020a<br>Mao et al., 2020 | 24/162 | 21/156 |
| Loss of Smell or Taste | Almazeedia et al., 2020<br>Mao et al., 2020<br>Lee et al., 2020 | 439/3823 | 47/337 |
| Abdominal Pain | Cholankeril et al., 2020<br>Zhang et al., 2020b<br>Mao et al., 2020 | 19/401 | 10/241 |
| Loss of Appetite | Zhang et al., 2020b<br>Mao et al., 2020 | 157/509 | 152/465 |
| Rash | Ratio from Galvan et al., 2020 applied to ZOE (2020) prevalence of 8.8%. | 7.8613/100 | 0.937/100 |

Quantitative data for ‘Other Conditions with COVID-like Symptoms’ conditioned on age and risk factor could not be readily found for the UK. Probabilities were estimated as follows: the ratio of deaths per age strata for respiratory disease in the UK, in 2012, was used to predict the prevalence in alive individuals (British Lung Foundation, 2013). Age categories were then further discretised, and prevalence was allocated as an exponential increase, as suggested by M. Xie *et al*., (2020). Finally, the estimates were categorised by high, medium and low risk, based on UK clinical risk ratios elicited from Cromer *et al*., (2014). The prevalence of influenza and COPD in the UK were discovered from Public Health England (2020) and Rayner *et al*., (2014) respectively.

### D. Testing

In the initial model, meningitis was included under Conditions with COVID-Like Symptoms, as a study by Packwood *et al*., (2020) reported a case of misdiagnosis. Meningitis can share several symptoms with COVID-19, namely fever, headache, lymphocytopenia and most indicatively, rash. However, it was omitted from the model as prevalence in the UK is low due to effective vaccination programs (McGill *et al*., 2020; PHE, 2019). COVID-19 infection has also been mistaken for dengue (Joob and Wiwanitkit, 2020), but it was not deemed appropriate for inclusion as dengue is not endemic to the UK.

AGENARISK supports sensitivity analysis via tornado diagrams. This allowed for the relative importance of each symptom in disease outcome to be compared. The analysis discovered cough, loss of appetite, shortness of breath, chest pain and chills to be the five most important contributors to severe COVID-19 infection (Fig. 3.). These symptoms differ, in part, to those recognised by the NHS (NHS, 2020). The difference may reveal significant symptoms that have previously been overlooked or imply a bias in the sample population dynamics. When applied to other conditions with COVID-like symptoms, the five most indictive symptoms were nausea or vomiting, cough, shortness of breath, abdominal pain and chest pain (Fig. 4.). Interestingly, the presence of anosmia or ageusia had the greatest negative impact on non-COVID-19 infection outcome, consistent with studies that express loss of sense of smell to be a marker for COVID-19 (Daher *et al*., 2020). As expected, elevated respiratory rate and reduced oxygen saturation had the highest relative importance to severe infection. Lymphocyte count had the lowest (Fig. 5.). For non-COVID infections, increased respiratory rate and elevated C-reactive protein were the most indicative laboratory findings (Fig. 6).

**Fig. 3.**
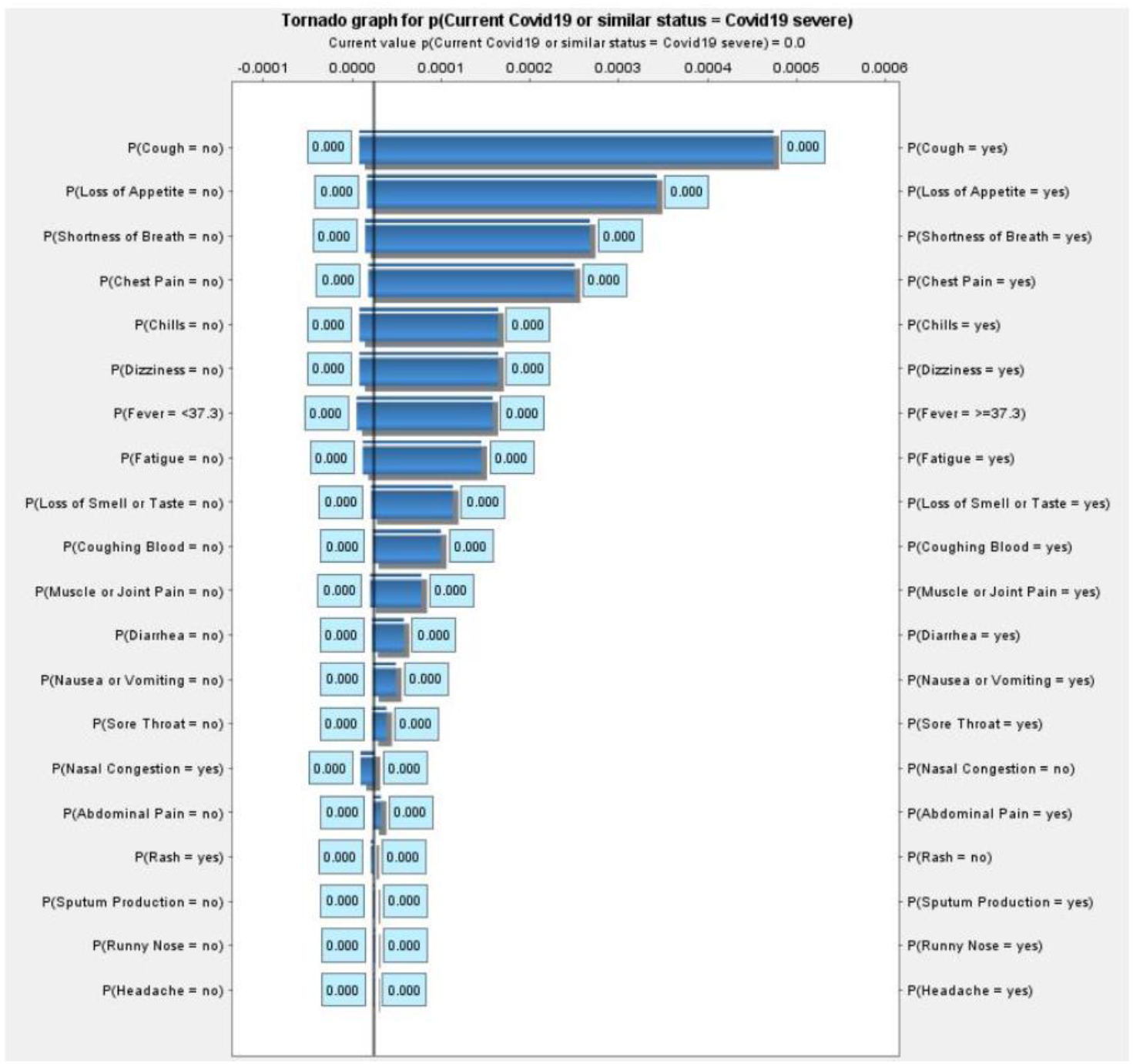
Tornado graph representing the hierachy of symptom sensitivities to severe COVID-19 infection.

**Fig. 4.**
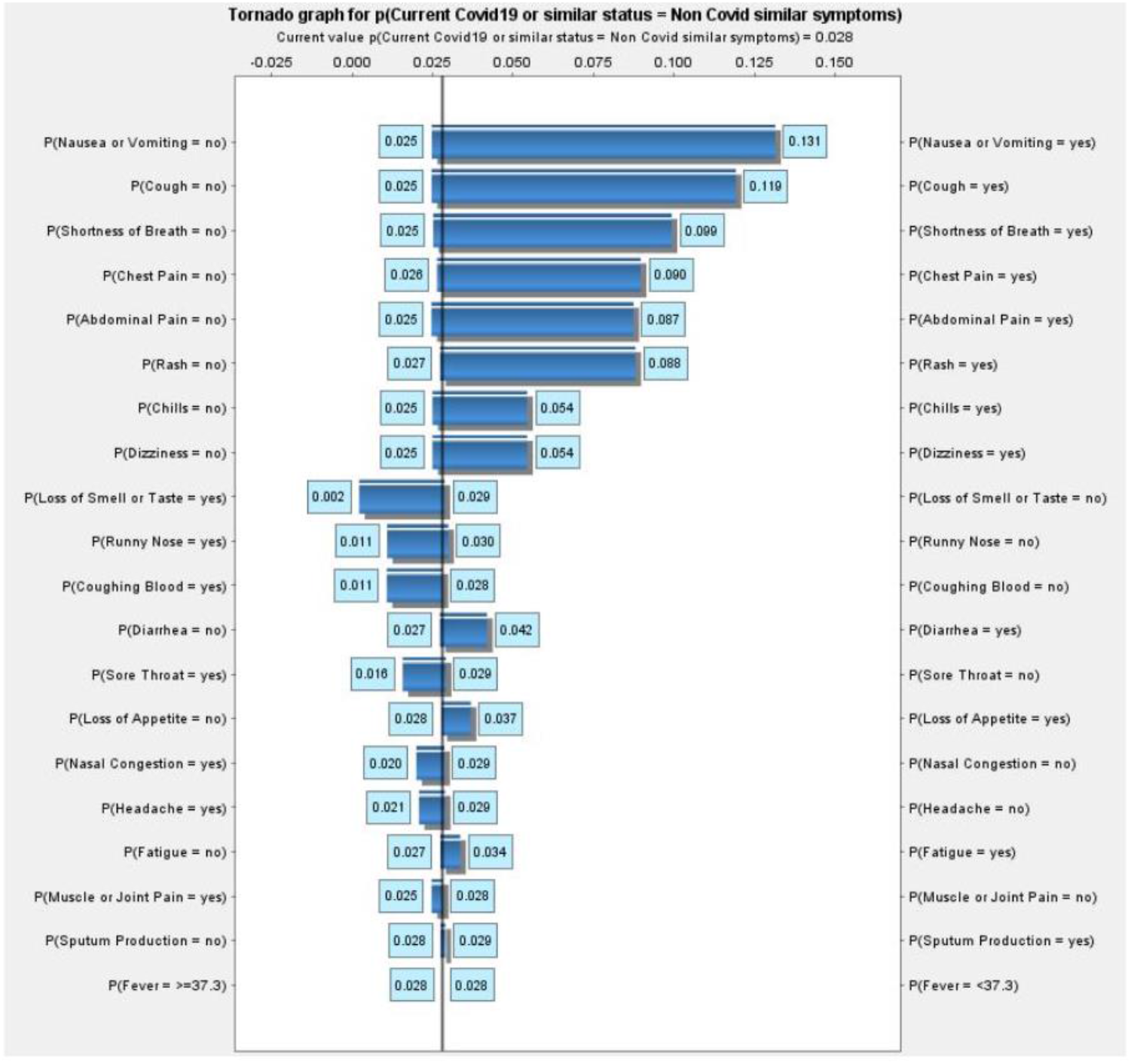
Tornado graph representing the hierachy of symptom sensitivities to non-COVID infection.

**Fig. 5.**
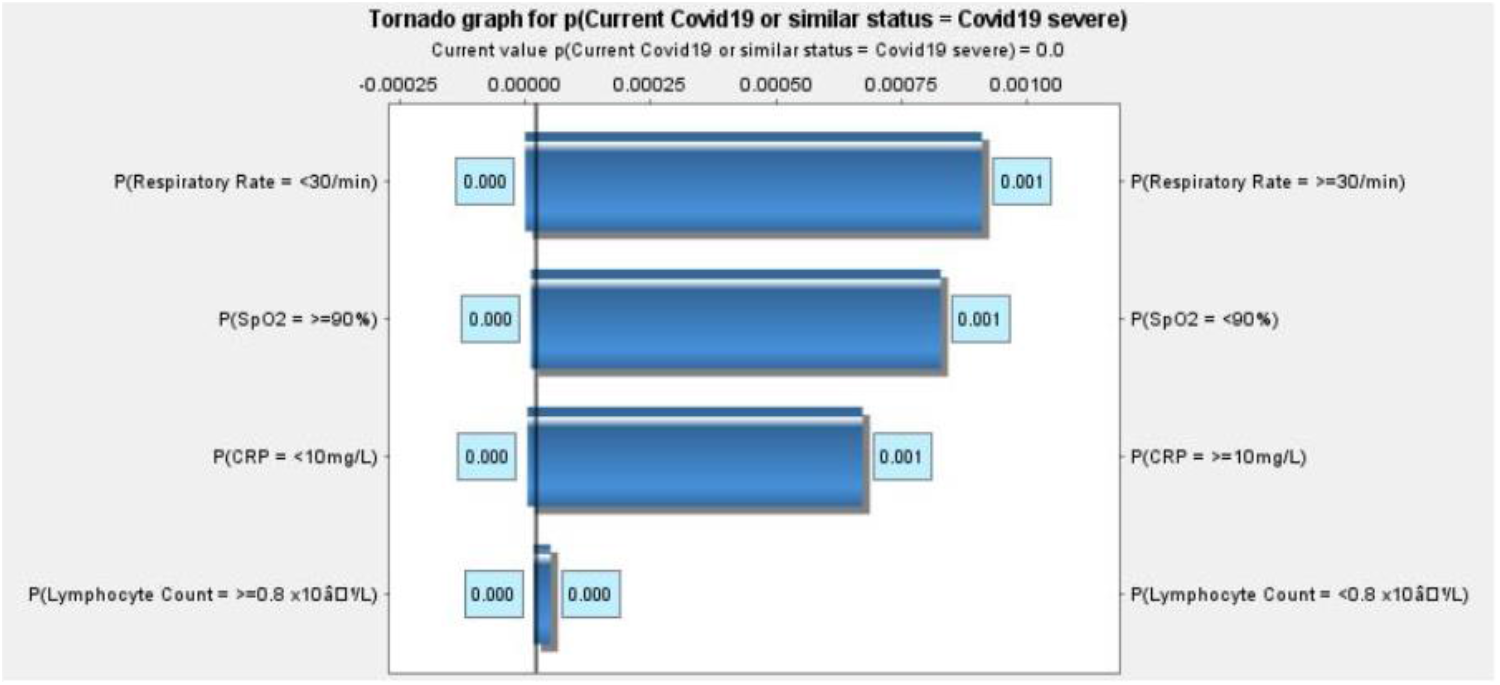
Tornado graph representing the hierachy of laboratory finding sensitivities to severe COVID-19 infection.

**Fig. 6.**
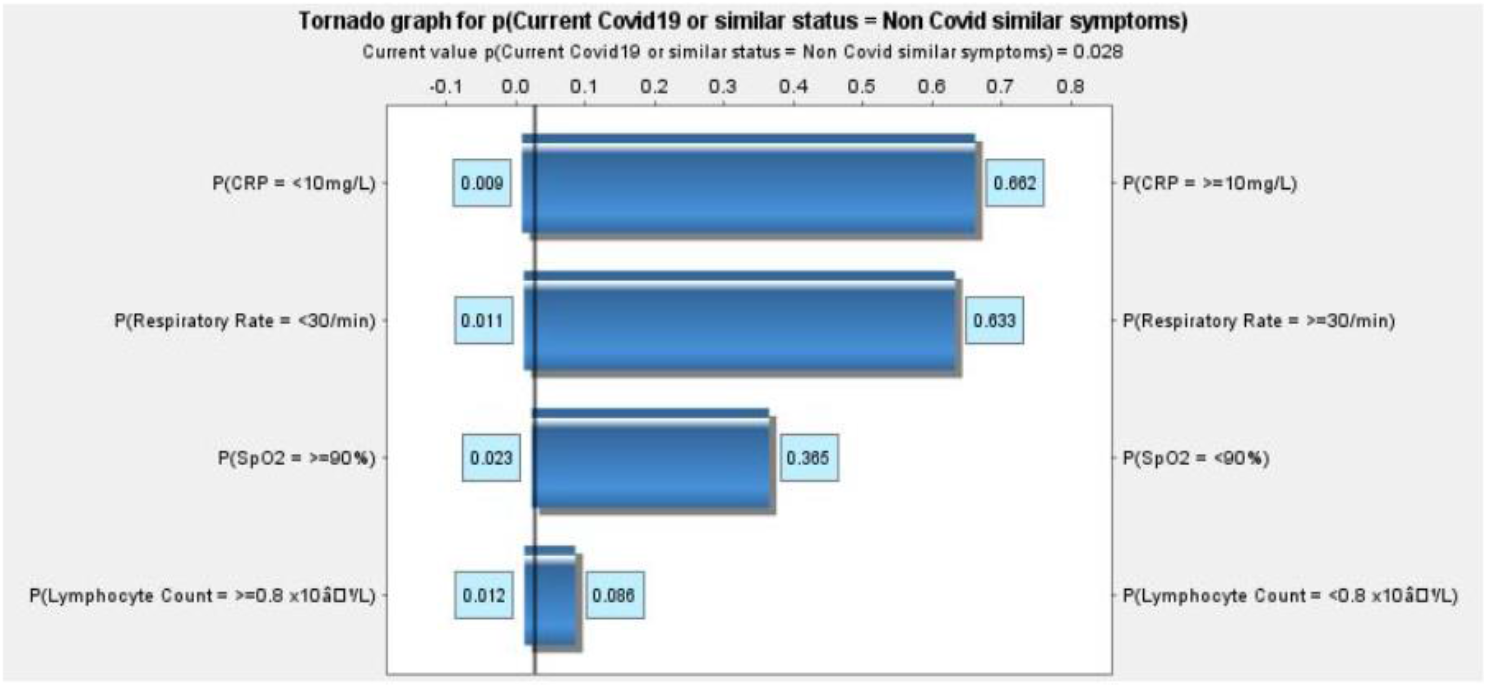
Tornado graph representing the hierachy of laboratory finding sensitivities to non-COVID infection.

## IV. Results

To test the model, subject profiles from case reports were input as scenarios. Study outcomes were then compared against the posterior probabilities of the network. All patients were positive for COVID-19. To assess the model’s ability to estimate infection, probabilities were compared before and after test node observations were added.

### A. Case Study 1

Sachdeva *et al*., (2020) reported a 71-year old, Caucasian female presenting with cough and deteriorating shortness of breath. The patient also complained of fever, but it was not clinically confirmed, and had no underlying medical conditions. Laboratory findings reported elevated C-reactive protein and normal lymphocyte count. She reportedly lived with an infected individual and was diagnosed with COVID-19 by a positive nasal PCR result. After treatment and respiratory recovery, the patient developed a maculo-papular rash. The nodes set in the network to reflect this can be found in Table. 4.

**TABLE IV.** Bayesian network scenario observations for case study 1.

| Node | Parameter |
| --- | --- |
| Ethnicity | White |
| Age | 65-74 |
| Sex | Female |
| Underlying Medical Conditions | Low |
| Contact with Confirmed Infected Person | Yes |
| Cough | Yes |
| Shortness of Breath | Yes |
| Symptom status | Deteriorating |
| CRP | $\geq 10\text{mg/L}$ |
| Test Type | Nasal PCR |
| Tested | Positive |

On initial run, the model predicted the patient to have a 49% probability of severe infection, and 43.85% probability of mild infection. Eventual status was also forecast to be severe (53.16%). With the prediction of severe infection and deteriorating symptom status, the ‘hospitalisation alert’ 60% threshold was breached (69.04%) (Fig. 7). With the addition of a positive RT-PCR observation, the primary diagnosis of severe infection was maintained for current (58.4%) and eventual status (62.26%). Mild infection was predicted at 40.64% (Fig. 8). To simulate the patient’s rash after respiratory recovery, observations for cough and shortness of breath were removed and rash symptom nodes were set to true for maculo-papular. It was also assumed that ‘current time since infected’ was >5 days and ‘symptom status’ was stable. No other observations were updated. Given these new parameters, our model overwhelmingly supported mild infection (93.47%) (Fig .9).

**Fig. 7.**
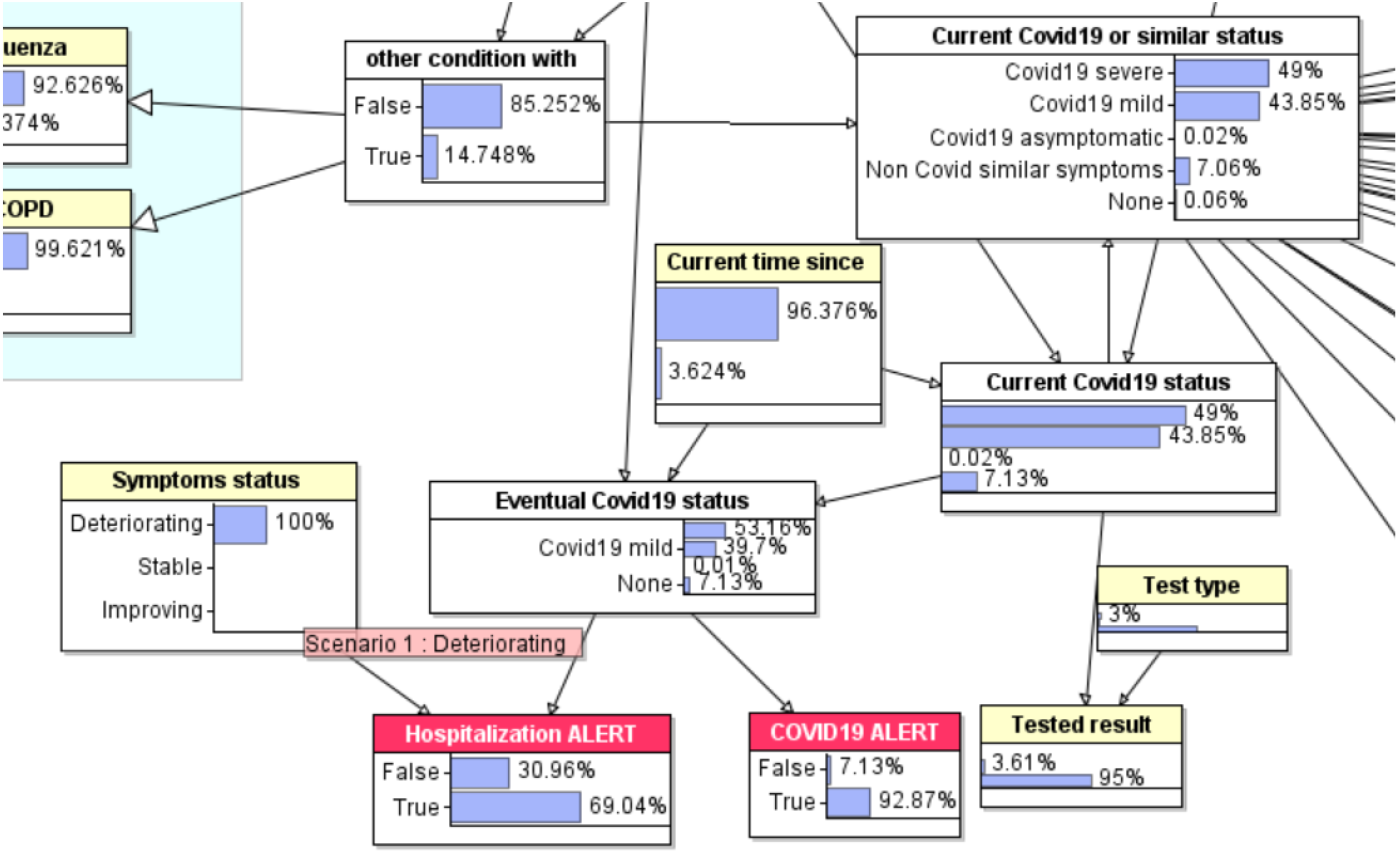
Posterior probabilities of COVID-19 status nodes for observations from case study 1, with ‘test type’ not yet observed.

**Fig. 8.**
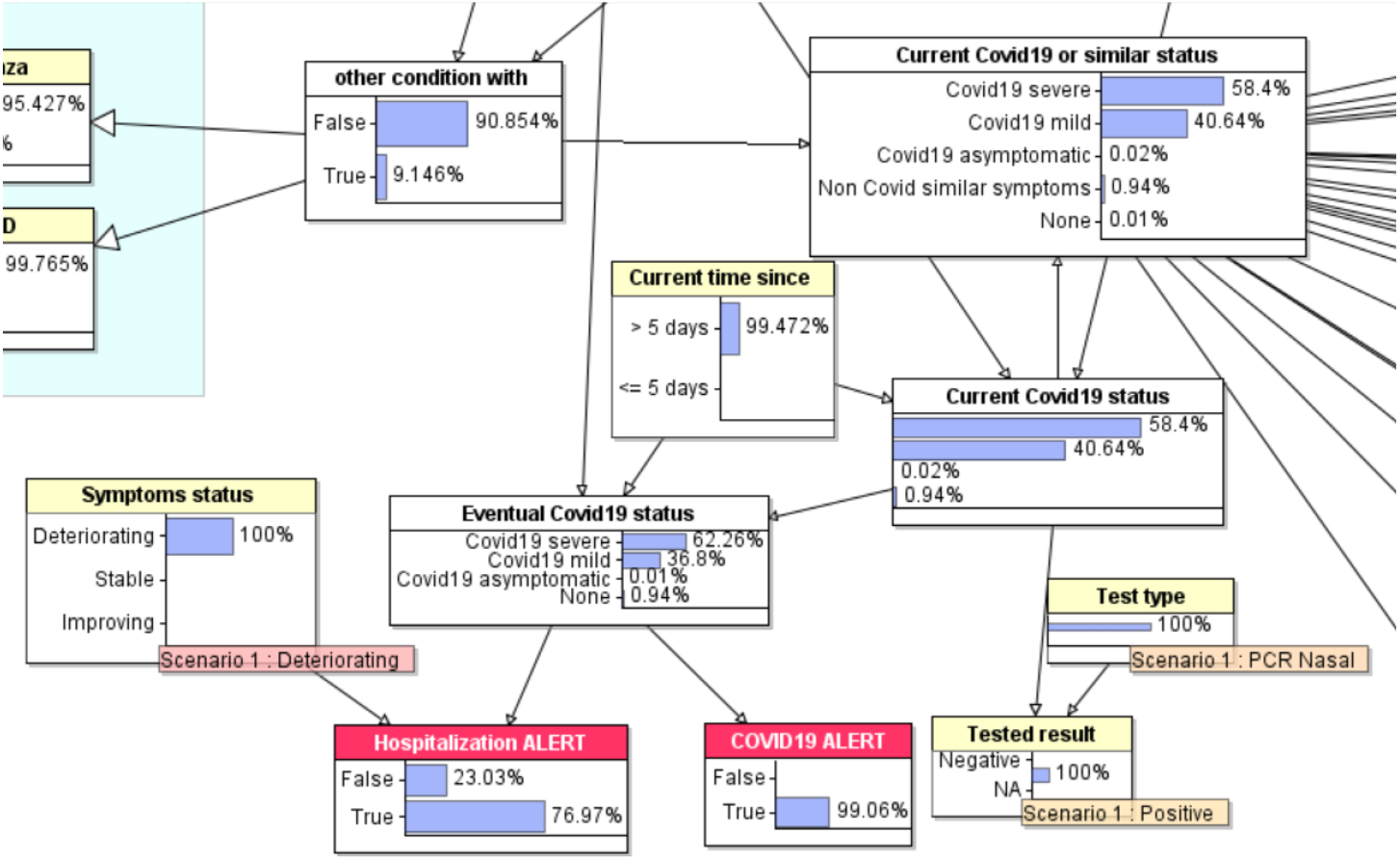
Posterior probabilities of COVID-19 status nodes for observations from case study 1, with ‘test type’ observed.

**Fig. 9.**
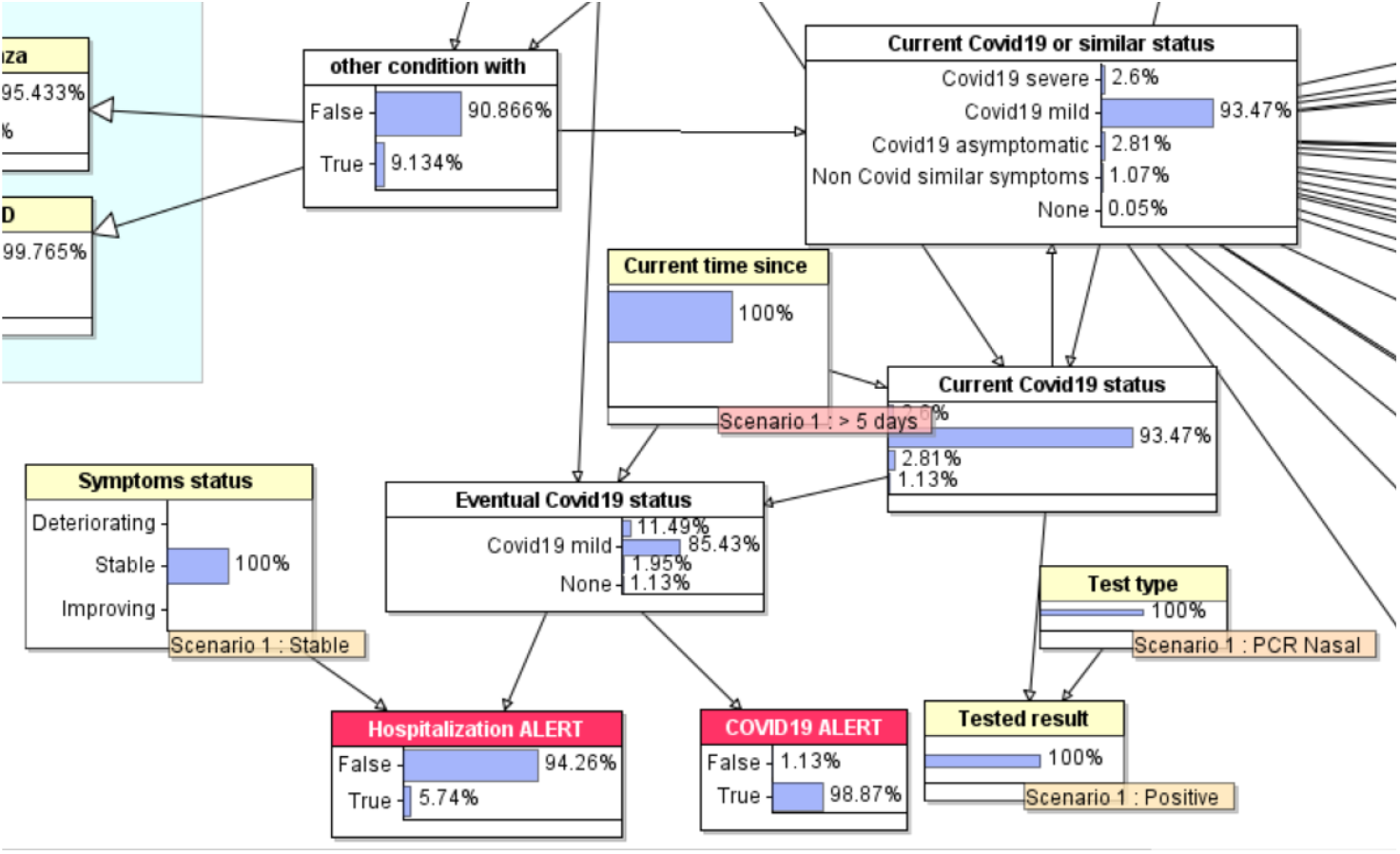
Posterior probabilities of COVID-19 status nodes after respiratory recovery and during rash presentation, for case study 1.

### B. Case Study 2

Chen *et al*., (2020c) reported a 46-year old, Chinese female presenting with a fever of 37.3°C progressing to sore throat, cough and chest pain after 5 days. The patient had an oxygen saturation of 98% and tested negative for influenza A and B. After admitting to having contact with a confirmed infected person, she was tested for SARS-CoV-2 via nasal RT-PCR and was confirmed positive. The nodes set in the network to reflect this can be found in Table. 5.

**TABLE V.** Bayesian network scenario observations for case study 2.

| Node | Parameter |
| --- | --- |
| Ethnicity | Chinese |
| Age | 35-54 |
| Sex | Female |
| Contact with Confirmed Infected Person | Yes |
| Fever | Yes |
| Sore Throat | Yes |
| Cough | yes |
| Chest Pain | Yes |
| Oxygen Saturation | $\geq 90\%$ |
| Test Type | Nasal PCR |
| Tested | Positive |
| Influenza | No |
| Current time since infected | $>5$ days |

Before confirming a positive test, our model strongly predicted mild infection at 83.16%, with a slight decrease in eventual status (77.2%). Severe infection was forecast at 16.57% for current and 22.53% for eventual status. As an observation was not available for symptom status, the prior probability favoured a stable progression and therefore ‘hospitalisation alert’ was not triggered (11.27%). ‘COVID-19 alert’ breached the threshold comfortably at 99.64%. After observing a positive PCR test, the probability of severe infection increased marginally to 20.38%, but mild infection was still the primary diagnosis (79.58%). The pattern followed for eventual status (26.09% and 73.87% respectively).

### C. Case Study 3

Song *et al*., (2020) reported a 37-year old, Chinese male comorbid with liver cancer. The patient began presenting with fever, cough and dyspnea, and was confirmed positive for influenza and SARS-CoV-2 via nasal PCR. He did not suffer from lymphocytopenia and observations were not available for C-reactive protein levels. The nodes set in the network to reflect this can be found in Table. 6.

**TABLE VI.** Bayesian network scenario observations for case study 3.

| Node | Parameter |
| --- | --- |
| Ethnicity | Chinese |
| Age | 35-54 |
| Sex | Male |
| Fever | Yes |
| Shortness of Breath | Yes |
| Cough | Yes |
| Influenza | Yes |
| Lymphocyte count | $\geq 0.8 \times 10^9/L$ |
| Test | Nasal PCR |
| Tested | Positive |
| Cancer | Yes |

**TABLE VII.** Bayesian network scenario observations for case study 4.

| Node | Parameter |
| --- | --- |
| Ethnicity | Chinese |
| Age | 0-15 |
| Sex | Male |
| Cough | Yes |
| Runny nose | Yes |
| Fever | No |
| Nausea or vomiting | No |
| Diarrhea | No |
| Contact with infected person | Yes |
| Lymphocyte count | $\geq 0.8 \times 10^9/L$ |
| CRP | $< 10 \text{mg/L}$ |
| Test | Nasal PCR |
| Tested | Positive |
| Current time since infection | $> 5$ days |

With the initial observations, the model predicted infection with a non-COVID-19 disease (99.51%) for both current and eventual status. Probability of mild and severe infection were almost inconsequential at 0.27% and 0.22% respectively. Given this scenario, neither ‘hospitalisation alert’ (0.13%) nor ‘COVID alert’ (0.49%) were triggered. With the addition of a positive RT-PCR result, infection with a non-COVID-19 disease was still favoured (96.25%). Severe COVID-19 was forecast at 1.95% progressing to an eventual status of 2.15%. Mild infection was predicted at 1.76% for current status and decreased to 1.6% for eventual. COVID-19 infection was also confirmed by a positive chest CT-scan. When this observation was entered, the probability of mild infection increased to 17.58% and severe infection to 15.05%. ‘Hospitalisation alert’ was not triggered for either test types. ‘COVID-19 alert’ threshold was not breached, despite positive COVID-19 test.

### D. Case Study 4

Li *et al*., (2020b) reported the infection of a 3-month old, Chinese male. The patient presented with cough and rhinorrhea but no fever, diarrhea or vomiting. Chest CT-scan suggested viral pneumonia. The child lived with a confirmed case and had been visited two weeks prior by relatives from Wuhan at the time of outbreak. Laboratory testing revealed elevated lymphocyte count and low CRP levels. COVID-19 infection was confirmed by positive RT-PCR. The nodes set in the network to reflect this can be found in Table. 7.

Initially the model strongly estimated ‘none’ for current status of COVID-19 infection at 78.39%. The probability of mild infection was calculated at 20.13% and severe at 1.2%. There was little change predicted in eventual COVID-19 status, with ‘none’ increasing to 78.41%, severe to 1.45% and mild decreasing to 19.94%. Neither ‘hospitalisation alert’ (0.73%) nor ‘COVID-19 alert’ (21.59%) were triggered. On the observation of a positive RT-PCR test, calculations favoured mild COVID-19 infection (60.88%). The probability of severe infection increased to 4.67% and ‘none’ to 33.87%. Eventual COVID-19 status followed a similar pattern (60.25%, 5.42% and 33.87% respectively). ‘Hospitalisation alert’ remained untriggered (2.71%) but ‘COVID-19 alert’ breached the threshold (66.13%).

## V. Discussion

For case study 1 the model correctly predicted severe COVID-19 infection. The patient was found to have pneumonia and required oxygen therapy, both indicators of severe disease. However, the prediction is weak with only 5.15% difference between mild and severe status before testing. This may be due to contradictory low risk health history and ethnicity with high risk age category and confirmed contact with infected persons. High-risk observations ultimately lead to a predicted increase in severe eventual status. However, the network does not account for the intervention of treatment on this node. Eventual status does not reflect the patient’s outcomes by the end of the study period, as she recovered fully after receiving antivirals and oxygen therapy. Treatment as a parent node of ‘eventual COVID-19 status’ could be a potential future development. The model was able to correctly trigger the ‘hospitalisation alert’ node. This prediction is concurrent with the study the patient was admitted to the Emergency Department. On the simulation of rash during convalescence, the update in prediction from 58.4% severe to 93.47% mild effectively reflects the patient’s recovery journey.

Mild COVID-19 severity was correctly predicted for the second case study. This is known from the study as treatment only consisted of antimicrobials and did not require additional ventilation. The observations did not trigger the hospitalisation alert, which may appear to contradict the report as the patient presented to the Third Affiliated Hospital of Sun Yat-sen University fever clinic. However, the case occurred during the early stages of the pandemic when clinical uncertainty was high. It is likely if the same case occurred at present day, hospitalisation would not have been necessary and hence the model would be correct.

For case study 3, the model evidently struggled with the non-specific symptom presentation and comorbidity observations. Despite the positive PCR results, a high probability was predicted for non-COVID-19 similar status. It is hypothesised that this occurred due to the model accounting for potential false-positives from the RT-PCR method (Tahamtan and Ardebili, 2020). Song *et al*., (2020) verified the initial diagnosis with a positive chest CT scan. With an observation of CT-scan for ‘test type’, the probability of mild infection increased, confirming the hypothesis. A potential development to the model could be the stratifying of ‘test type’ such that PCR and CT-scan are not mutually exclusive and can be observed concurrently. Even with the updated test type, probabilities were not as explicit as hoped in comparison to the outcome of the study. The patient was admitted to the ICU and administered oxygen, which convincingly points to severe infection, whereas our model predicted a non-COVID-19 disease. As the patient had recently undergone chemotherapy, he may have been temporarily at higher risk for severe infection due to the immunocompromising cancer treatment (Williams et al., 2020b). It could be argued that if the patient had not recently undergone chemotherapy, his disease severity would have been mild, supporting the model outcome. However, research shows this hypothesis is a point of contention, with studies such as Jee *et al*., (2020) disputing chemotherapy as a risk for COVID-19 severity.

The subject of case study 4 was predicted to be negative for COVID-19 infection. This is understandable as less than 5% of COVID-19 infections reported in the EU/EEA and UK are in children under 18 years old (ECDC, 2020b). Similarly, many of the subject’s observations denied the presence of COVID-19. For example, diarrhea, vomiting, fever and laboratory findings were actively unobserved in the patient, lowering the probability of infection. Bias in the data used to populate the model was often unavoidable and therefore must be considered. Most data available for COVID-19 infection are from adults. Similarly, symptoms conditioned on age were not explored. When a positive RT-PCR test was observed, the model was able to accurately diagnosis the subject with mild COVID-19. This is concurrent with the study as recovery only required symptomatic treatments, such as cough medicine and traditional Chinese tonics (Li *et al*., 2020b).

## VI. Conclusion

As the future of the COVID-19 pandemic remains uncertain, governments and researchers push to develop technologies and strategies to regain control. This paper proposes a Bayesian network to aid in diagnosis and triage of potential COVID-19 positive individuals. The model demonstrated high efficacy in predicting the disease outcomes of initial case studies but should be subject to larger raw datasets for more rigorous testing. One of the main achievements is the network is not reliant on the observation of a clinical test result for accurate prediction. This has huge potential benefits in monitoring community prevalence, especially in populations where clinical testing is inaccessible. Distinguishing COVID-19 from diseases with similar presentations is identified as an area for improvement and may prove a caveat as we transition into winter influenza season. Treatment as an intervention on ‘eventual COVID-19 status’ and the separation of ‘test type’ to allow for mutual inclusivity should be considered for future developments. Going forward, it is hoped this model can be implemented in a tangible setting, for contribution to COVID-19 relief efforts.

## VII. Future Work

Governments internationally are increasingly turning to novel digital technologies in response to the pandemic. Contact tracing is a well-established protocol in infectious disease management that functions to disrupt transmission among population clusters. The concept involves individuals maintaining a log of recent contacts and locations. If they or a person in their network contracts the disease, the likely transmission route can be traced and appropriate action taken (Braithwaite *et al*., 2020). Developers are leveraging ubiquitous personal devices, such as smartphones, to efficiently reach the wider population. Across governments, app-based contact tracing for COVID-19 is a growing craze. Whilst England and Scotland are amid developments of their respective test and trace applications, Northern Ireland has already released the StopCOVID NI Proximity App. The app supports a decentralised framework in which users’ smartphones exchange digital keys, via Bluetooth, if within a two-metre vicinity for longer than fifteen minutes. When a user informs the app of a positive test result, keys collected from the previous fourteen days will be used to alert their respective owners. In line with government guidelines, alerted users must self-isolate for two weeks (nidirect, 2020).

A caveat to this approach is a positive clinical test must be obtained for input into the application. The model developed in this paper could replace the need for a test with a threshold probability score or triggering of the ‘COVID alert’ node. The Bayesian network could be embedded in the application and interact with a graphical user interface via the AGENARISK application programming interface (API). In the interest of data privacy, only the user’s location and disease probability score would be forwarded to the government for monitoring. This application is currently in development by Fenton *et al*., (2020).

## Data Availability

The authors confirm that the data supporting the findings of this study are available within the article and its supplementary materials.

## Acknowledgment

I would like to thank my supervisor Norman Fenton for his expert guidance and for kindly providing the license to AGENARISK for the duration of this project. I would also like to thank the group, Norman Fenton, Scott McLachlan, Peter Lucas, Kudakwashe Dube, Graham Hitman, Magda Osman, Evangelia Kyrimi and Martin Neil, whose research this project builds upon. Finally, I would like to thank fellow student Georgina Prodhan for her contributions to the model.

## Appendix

